# Global Menstrual Cycle Symptomatology as Reported by Users of a Menstrual Tracking Mobile Application

**DOI:** 10.1101/2022.10.20.22280407

**Authors:** Alessandra J Ainsworth, Kimberly Peven, Ryan Bamford, Liudmila Zhaunova, Rodion Salimgaraev, Carley Prentice, Aidan Wickham, Adam Cunningham, Frederick Goddard, Sonia Ponzo, Samir N Babayev

## Abstract

**Background:** Although a shared experience throughout the world, menstrual symptomatology is underreported and often misunderstood, by both individuals who menstruate and their providers. This study aimed to investigate symptomatology throughout the menstrual cycle, including the frequency of symptoms reported for each phase of the cycle and the relationship between symptoms.

**Methods:** This study included self-reported cycle information and symptoms during ovulatory menstrual cycles in mobile application users. Symptom frequency was described overall, by phase, and by day. Relationships between symptoms were examined through correlation coefficients and term frequency-inverse document frequency analysis.

**Results:** 437,577 unique users and 896,051 unique cycles were included in the analysis. Somatic symptoms were most frequently reported, logged in 88.3% of cycles. Of the total symptoms logged, somatic, gastrointestinal, and negative mood symptoms were most prevalent in the late luteal and bleeding days. In contrast, positive mood and discharge were most reported in the fertile window. Mood symptoms were highly correlated with each other (energetic mood and happy mood r=0.62, anxious mood and obsessive thoughts r=0.60). Fatigue was highly correlated with somatic and gastrointestinal symptoms such as bloating (r=0.62), headache (r=0.60), backache (r=0.58), and nausea (r=0.55).

**Conclusions:** Menstrual cycle-related symptoms are very common and vary throughout the menstrual cycle. While negative symptoms, including pain, bloating, and negative mood, are most commonly reported, women also report positive symptoms, such as energetic and happy mood. Self-reported data from cycle tracking and health apps can improve our understanding of the prevalence and variability of menstrual-related symptoms.

**Plain English Summary:** Our study includes data collected from users of a menstrual tracking application. We found that menstrual symtpoms are common, throughout the menstrual cycle, and include both positive and negative mood and physical symptoms. This large data set may help to normalize menstrual symptoms and provide a basis for future research and understanding.

## Background

Although a shared experience by women and people who menstruate throughout the world, menstruation represents an area of health often ignored and misunderstood by both patients and healthcare providers. While many individuals experience menstruation without significant symptoms, others endure pain and many other somatic, gastrointestinal, and mood symptoms which vary throughout the cycle. Dysmenorrhea affects 45-95% of menstruating individuals, with wide variation due to differing definitions of the condition (1). Compared with pain-free phases of their cycles, those suffering from dysmenorrhea experience reduced quality of life, poor mood, and decreased sleep during menstruation (1). Gastrointestinal symptoms also vary throughout the menstrual cycle, with one-third of women experiencing symptoms during menstruation (2), most frequently on the first day of bleeding (3).

Additionally, irritability and mood swings can affect 22-40% of women, more often before menstruation (4). As it relates to symptom co-occurrence, individuals with mood symptoms like depression and anxiety are more likely to experience multiple gastrointestinal symptoms both before and during menstruation (5). Despite improved standardization in terminology and increasing prioritization of women’s health issues, these symptoms are under reported (6–8). Importantly, menstrual symptoms may present a significant impairment to productivity and quality of life (9).

A recent review highlighted challenges in the field of menstrual health research (10). These challenges include how and from whom data is collected, normalization of symptoms, stigma regarding menstrual disorders, lack of standardized assessment tools, and differing cultural perceptions of this physiologic process (10–13). Previous studies of symptom prevalence and variation throughout the menstrual cycle are small prospective or survey studies (4,5). Given these limitations, there remain gaps in our knowledge of patient symptoms and opportunities for further study.

The emerging availability of mobile applications (apps) and user-reported data provides a novel approach to population health, in this instance, menstrual symptomatology, through ‘digital epidemiology’ (14–16). The impact of digital epidemiology to inform our understanding of population norms in menstrual health recently demonstrated a luteal phase length of 12.4 ± 2 days (17). These results differ from the classical teaching of a conserved 14-day luteal phase (17).

In this study, through in-app user-reported data, we aim to report a population-based assessment of symptomatology throughout the menstrual cycle. Specific objectives include describing the frequency of symptoms reported in each phase of the menstrual cycle and analysis of the relationship between symptoms.

## Methods

This cohort study includes active users of the Flo app recording symptoms during an ovulatory cycle starting between January 1, 2021, and January 1, 2023. Users aged 18-45, not pregnant, and actively involved in the app (reporting at least three menstrual cycles) were included. Menstrual cycles between 21-35 days, menstrual bleeding between 1-10 days, follicular and luteal phase lengths of at least five days each, positive ovulation predictor kit use, and at least one self-reported menstrual symptom at any point in the cycle were included as ‘normal’ menstrual cycles.

Cycle dates and phases were calculated based on user-provided data. Users report the start and end dates of their period and the date of positive ovulation test. Each cycle start date is defined as the first day of bleeding. The mid-follicular phase was defined as the day after the last day of menstrual bleeding until the start of the fertile window (Figure 1). An individual with less than five days between the end of menstrual bleeding and the day of ovulation had no defined mid-follicular phase. The fertile window was calculated as the five days preceding ovulation and the day of ovulation was estimated as the day following a user-reported positive ovulation predictor kit. The luteal phase was split evenly in half and described as the early luteal phase and the late luteal phase.

Symptoms were self-reported and associated with the phase of the menstrual cycle as defined above. We classified symptoms into five groups: somatic (cramps, tender breasts, headache, fatigue, backache, acne), gastrointestinal (GI) (bloating, nausea, diarrhea, cravings, constipation), positive mood (happy, high sex drive, energetic), negative mood (mood swings, anxious, depressed, insomnia, stress, obsessive thoughts), and discharge (creamy, eggwhite, sticky, watery). Users could record as many symptoms in each category as desired each day.

Individual characteristics were matched to each cycle. Age at the time of the cycle was based on the user’s birthdate and cycle dates. A user’s country was assigned based on phone settings at app installation. Where the country at app installation was unavailable, country phone settings at the last login was used. Country prevalence of symptoms was evaluated only when 100 users or more were included.

## Statistical Methods

Using a cycle-level unit of analysis, we employed descriptive statistics to describe cycle length and symptoms experienced overall and by phase. For cycles with a length of 25-30 days, we describe symptoms experienced by day of the cycle, where day 1 is the first day of bleeding. We examined the correlation between all possible pairs of symptoms using a cycle-level unit of analysis considering symptoms experienced at any point in the cycle. We reported Pearson correlation coefficients for each possible pair. We also conducted a term frequency-inverse document frequency (tf-idf) analysis (18) for pairs of symptoms separately by cycle phase to describe the frequency of pairs of any symptoms, taking into account the frequency of the individual symptoms that make up the pair. This is calculated as term frequency times inverse document frequency, where term frequency is defined as the number of instances of a pair of symptoms in a cycle phase, and inverse document frequency is defined as the logarithm of the number of users logging one symptom from the pair during the relevant cycle phase divided by the total number of symptoms logged in the phase. Data were analyzed in Python Jupyter Notebook 6.0.1 and R version 4.2.0.

## Results

A total of 437,577 unique users and 896,051 unique, normal menstrual cycles met inclusion criteria with an average of 2.05 ± 1.8 cycles per user (Figure 2). The mean age of users was 30.0 ± 5.4 years; users under age 18 or over 45 were excluded from the analysis. Users were included from 222 countries, with 113 countries having at least 100 users. The largest group of users was in the United States (n=126,352), followed by Germany (n=45,018) and the United Kingdom (n=35,007).

Of the 437,577 unique users included in this analysis (last cycle), the mean cycle length was 28.4 ± 2.6 days (median 28.0 days), and the mean length of bleeding was 4.0 ± 1.1 days (median 4.0 days). Most included cycles had a length of 25-31 days (75.7%, n =331,360), followed by a length of ≥32 days (15.8%, n=69,479) and <25 days (8.4 %, n =36,738). Calculated from the predicted day of ovulation, the mean follicular phase length amongst users was 14.4 ± 3.0 days (median 14.0 days), and the mean luteal phase length was 14.0 ± 2.7 days (median 14.0 days). Of the 896,051 unique cycles, somatic symptoms were reported most often; 88.3% (n=790,923) of cycles reported at least one somatic symptom. Comparatively, gastrointestinal symptoms and discharge symptoms were reported in 60.8% of cycles each (n=545,107 and 544,460, respectively), negative mood symptoms were reported in 43.3% (n=388,249) of cycles, and positive mood symptoms were reported in 35.4% (n=317,558). The type and frequency of symptoms are shown in Figure 3. Menstrual cramps, breast tenderness, headache, and fatigue were the most common symptoms reported.

Symptoms were next assessed by the menstrual cycle phase. Somatic, gastrointestinal, and negative mood symptoms were most frequent in the late luteal phase and least frequent in the mid-follicular phase (Figure 4). 57.3% of all logged instances of tender breasts occurred during the late luteal phase compared to 1.8% in the mid-follicular phase. Discharge and positive mood symptoms were most common during the fertile window. 63.1% of all logged instances of egg white discharge and 45.3% of all logged instances of high sex drive were during the fertile window.

For cycles between 25-30 days in length (n=570,123 cycles), we examined daily symptom logs for the three symptoms with the most variability per category (Figure 5). Overall, most symptoms are reported on day 1 of the cycle (436,812 symptoms logged from 211,021 cycles out of 570,123 total cycles); however, the fertile window is where most cycles have at least one symptom logged. Out of 570,123 total cycles, 382,115 cycles had at least one symptom logged during the fertile window, and 283,652 had at least one symptom logged during the bleeding days. The prevalence of somatic, gastrointestinal, and negative mood symptoms are highest on day 1 but decline quickly, reaching their lowest point around day 7 before slowly increasing throughout the rest of the cycle. For positive mood symptoms, the prevalence of energetic mood remains relatively stable throughout the cycle. In contrast, happy and high-sex-drive moods increase slowly in the early part of the cycle, peaking around day 14 of the cycle and reaching baseline around day 20. Discharge symptoms are rarely logged during the early part of the cycle where bleeding occurs. Egg white discharge increases rapidly between days 6 and 13 before decreasing quickly to a low level by day 20.

For countries with at least 100 users, the number per country ranged from 103 in The Bahamas to 126,352 in the United States (Supplemental Table 1). The prevalence of cramps during the last cycle ranged from 30.5% in Vietnam to 74.6% in Iceland (Figure 6). Bloating ranged from 17.6 in Thailand to 49.7% in Chile. Mood swings ranged from 13.3% in Hong Kong to 43.1% in Iraq.

Mood symptoms were most highly correlated with each other. The anxious mood was highly correlated with obsessive thoughts (r=0.60) and depressed mood (r=0.58). Depressed mood was correlated with obsessive thoughts (r=0.55), and happy mood was highly correlated with energetic mood (r=0.62, Figure 7). Similarly, fatigue was highly correlated with somatic and gastrointestinal symptoms such as bloating (r=0.62), headache (r=0.60), backache (r=0.58), and nausea (r=0.55).

A term frequency-inverse document frequency (tf-idf) analysis by cycle phase showed how pairs of symptoms correlate within cycle phases, adjusting for the frequency of individual symptoms (Figure 8 and Supplementary Tables 2 and 3). During the bleeding days and late luteal phase when cramps are common, the top five tf-idf values all included cramps, and cramps were most closely associated with nausea (tf-idf = 54,861 bleeding days, tf-idf = 68,467 late luteal). Happy and energetic were the top pair in the mid-follicular phase (tf-idf=23,333) and second in the fertile window (tf-idf=55,636).

## Discussion

In this study, we report the frequency of somatic, gastrointestinal, mood, and discharge symptoms experienced throughout the menstrual cycle by a cohort of global, ovulating period-tracking app users. Notably, most users were aged 30-34 and located in Europe and the United States. Our results build on prior work evaluating the menstrual cycle characteristics of women using period-tracking apps. The median cycle length, 28.8 days in our study, is consistent with previous studies and period tracking app data (17,19–22).

Somatic symptoms were reported most frequently, with cramps being the most reported symptom. Somatic, gastrointestinal, and negative mood symptoms were more common in the late luteal phase than in the rest of the cycle. They reached a nadir in the mid-follicular phase between bleeding and the fertile window/ovulation. The hormonal changes observed in the luteal phase may explain these findings. There is a direct correlation between the rapid rise of progesterone, the dominant sex hormone in the luteal phase, and the abrupt increase in somatic symptoms. Progesterone has been shown to influence breast changes and associated breast tenderness in the luteal phase (23). By thermoregulatory actions, progesterone has also been shown to impact circadian rhythms leading to decreased rapid eye movement (REM) sleep in the luteal phase and, thus a reported increase in fatigue (24). However, progesterone levels fall in the week prior to menstruation while somatic symptoms continue to increase. Symptoms in the late luteal phase reflect the effect of decreasing systemic sex steroid hormones. Pain disorders, such as migraines and non-migraine headaches, have been shown to increase in the late luteal phase when estradiol levels are lowest (25–27), in line with our findings of headaches being most reported in this phase. These effects are modulated by estrogen effects on central pain processing networks and cerebral vasculature (26). Conversely, decreasing progesterone levels and the resultant increase in available androgen receptors may explain the increase in acne reported in the late luteal phase (28).

Gastrointestinal symptoms that increase throughout the luteal phase likely reflect multiple mechanisms (3). First, the effect of progesterone on serotonergic activity in both the central nervous system and the GI tract slows gastrointestinal motility leading to constipation in the early luteal phase (29). Second, increasing prostaglandin levels in the late luteal phase lead to stimulation of smooth muscle contractions – leading to both uterine cramping and increased gastrointestinal motility and associated diarrhea (30).

The effect of the menstrual cycle on mental health has been well studied, and premenstrual mood disorders are well classified, with over 300 subtypes reported. Mood changes in the luteal phase affect 50-80% of reproductive-aged women, although 3-8% of individuals have such severe symptoms with associated functional impairments that they meet diagnostic criteria in the DSM-V (31). Our study confirmed an increase in negative mood symptoms throughout the luteal phase, possibly reflecting known alterations in sex steroids and the following neurotransmitters: serotonin, noradrenaline, and gamma-aminobutyric acid (GABA) (31,32). Negative mood symptoms decreased throughout the follicular phase, with a corresponding increase in positive mood symptoms that reached peak levels in the late follicular phase. Positive mood symptom frequency in the follicular phase is consistent with prior studies reporting individual well-being to be highest at this time in the cycle (33,34). Recent work utilizing functional magnetic resonance imaging (fMRI) has added to our understanding. Sex hormones appear to impact communication between brain regions, in addition to their known impact on neurotransmitter secretion (35). This impacts cognition and emotional processing, such that estrogen seems to decrease reaction to negative stimuli while progesterone seems to increase emotional reactions to negative stimuli (36).

The relationship between mood and somatic symptoms during the menstrual cycle has been evaluated with mixed results. One study found an increase in mood symptoms in women with gastrointestinal symptoms, skin changes, breast changes, and abdominal bloating. At the same time, there was no effect of concurrent mood symptoms seen in women who reported back and joint pain (37). We found negative mood symptoms (depressed mood, mood swings, anxious mood) co-occurred with cramps in the bleeding days and late luteal phase while positive mood symptoms (energetic and happy) occurred together in the mid-follicular through early luteal phases, without strong associations with somatic symptoms.

We found small geographic differences in user-reported symptoms during the menstrual cycle. This may reflect the impact of knowledge or sociocultural constructs, which impact individual perceptions of symptoms and their menstrual experience (38). Historically, menstruation has been described negatively. Previous work has shown an association between early menstrual/menarchal experiences and current menstrual experience, body perception, and overall health (39–42). However, other studies examining symptom experience across broad geographic regions have not found regional differences in symptoms (43).

The strength of this study lies in the number of unique users and in the large amount of user-reported data from a global population. However, this study is not without limitations. Specifically, this study is subject to selection bias as not all menstruating individuals choose to use period-tracking apps. Demographic and other individual characteristic data were not collected, including user-reported race and ethnicity, which may be important mediators of menstrual symptom characteristics and experiences that may impact our results. We included only ovulatory cycles where ovulation was confirmed by self-ovulation test to more accurately divide the cycle into phases. Users logging ovulation tests likely have a goal of conception and may thus have a different experience of menstrual symptoms and a difference in the tendency of logging symptoms. Additionally, irregular and non-ovulatory cycles are not represented in this analysis, and results are not generalizable to these populations. Although this study represents a global menstruating population, many countries remain absent or underrepresented, presenting an opportunity for future research. Other limitations include the use of self-reported symptom-tracking data. Users log symptoms through the app but may not log symptoms consistently throughout the cycle (e.g., potentially more likely to log symptoms when logging other events such as ovulation or period) and may not log all symptoms experienced. Users also may be more likely to log symptoms that appear at the top and front of the app over symptoms that appear lower down or require scrolling to see. This decreases the representativeness of symptoms recorded and limits our ability to compare results with other methods of symptom data collection.

## Conclusions

Our study presents an expanded understanding of symptoms experienced during the menstrual cycle. These findings reflect the value of population-level menstrual cycle data, which can help inform individual patients and clinicians while strengthening research efforts and calls for advocacy for Women’s Health.

## Abbreviations

GI: gastrointestinal
Tf-idf: term frequency-inverse document frequency
REM: rapid eye movement
DSM-V: Diagnostic and Statistical Manual of Mental Disorders, 5th edition
GABA: gamma-aminobutyric acid
fMRI: functional magnetic resonance imaging

## Declarations

1. IRB approval waived. Documentation attached as supporting document
2. Consent for publication: Not applicable
3. Availability of data and materials: The datasets used and/or analyzed during the current study are available from the corresponding author on reasonable request.
4. Competing interests: KP: Flo Health UK Limited (Employment, ownership interest, stock/stock options) RB: Flo Health UK Limited (Employment) LZ: Flo Health UK Limited (Employment, ownership interest, stock/stock options) RS: Flo Health UK Limited (Employment, ownership interest, stock/stock options) CP: Flo Health UK Limited (Employment) AW: Flo Health UK Limited (Employment, ownership interest, stock/stock options) AC: Flo Health UK Limited (Employment) FG: Flo Health UK Limited (Employment) SP: Flo Health UK Limited (Employment, ownership interest, stock/stock options). Other authors did not report any potential conflicts of interest.
5. Author contributions: AJA: Design, interpretation of data, drafted and substantially revised work KP: Design; acquisition, analysis, and interpretation of data; drafted and substantially revised work RB: Acquisition, analysis, and interpretation of data LZ: Design, interpretation of data, and substantively revised work RS: Design; acquisition, analysis, and interpretation of data CP: Analysis and interpretation of data AW: Acquisition, analysis, and interpretation of data AC: Acquisition, analysis, and interpretation of data FG: Analysis and interpretation of data SP: Interpretation of data and substantively revised work SB: Design, interpretation of data, substantially revised work
6. Acknowledgements: None

## Data Availability

All data produced in the present study are available upon reasonable request to the authors.

---

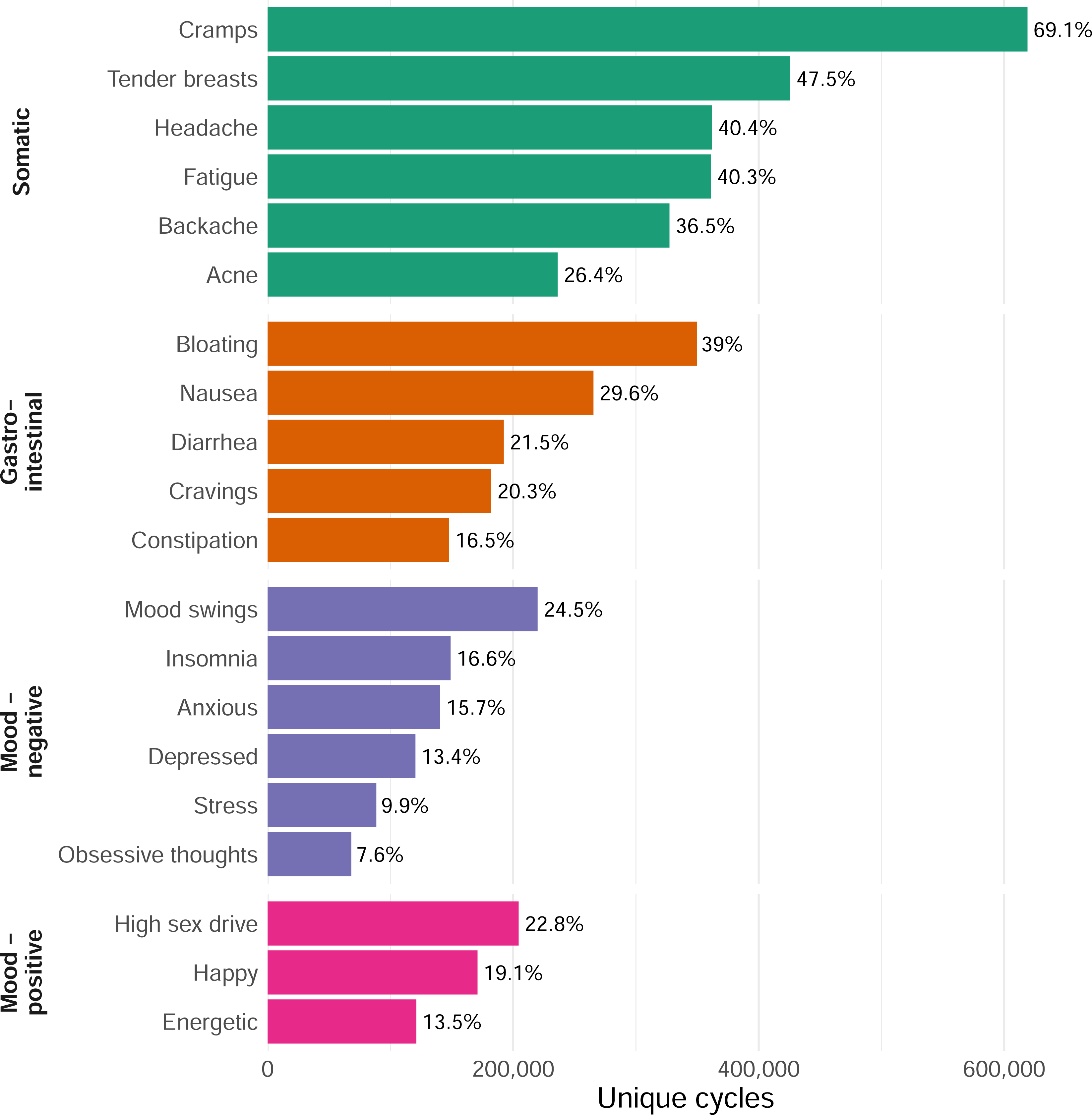

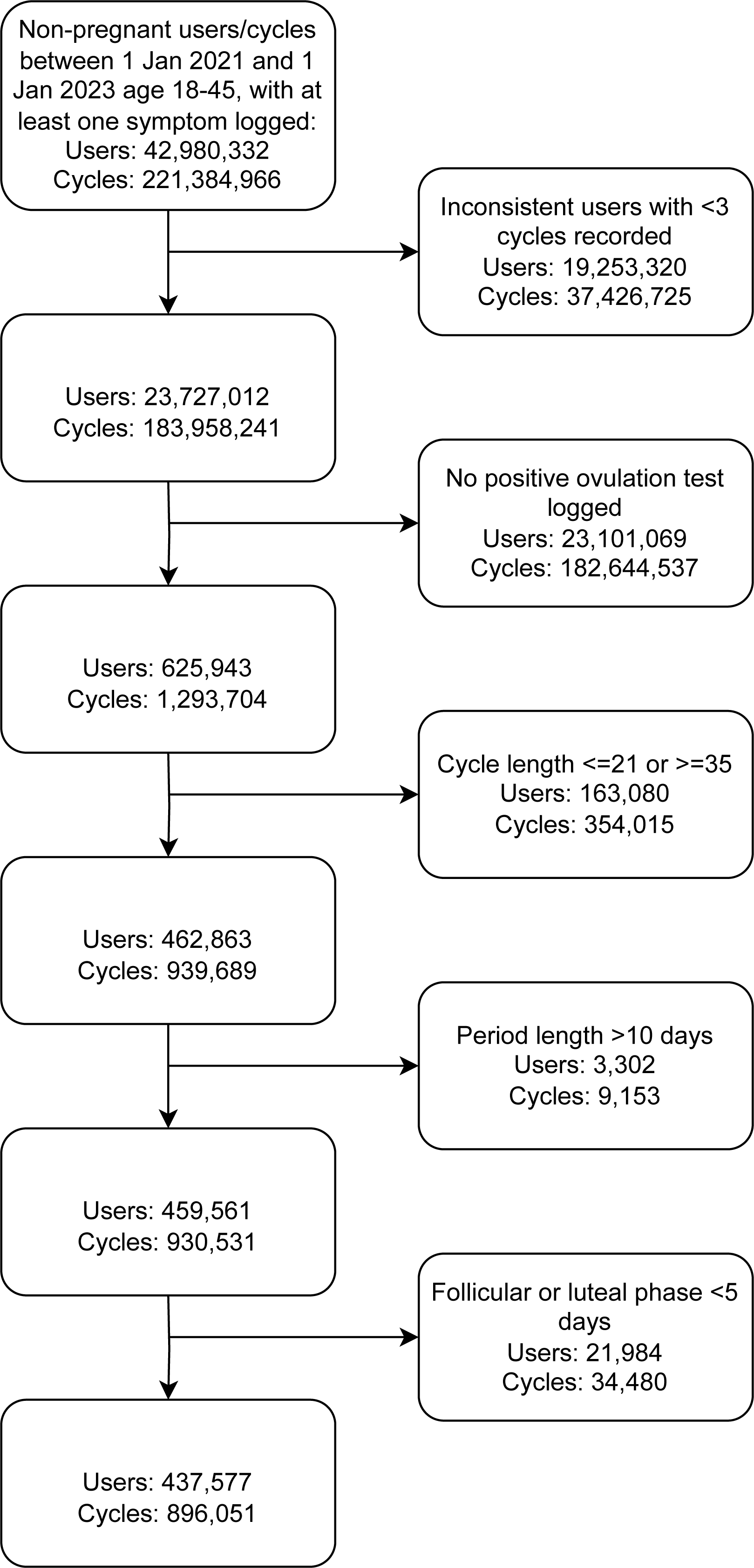

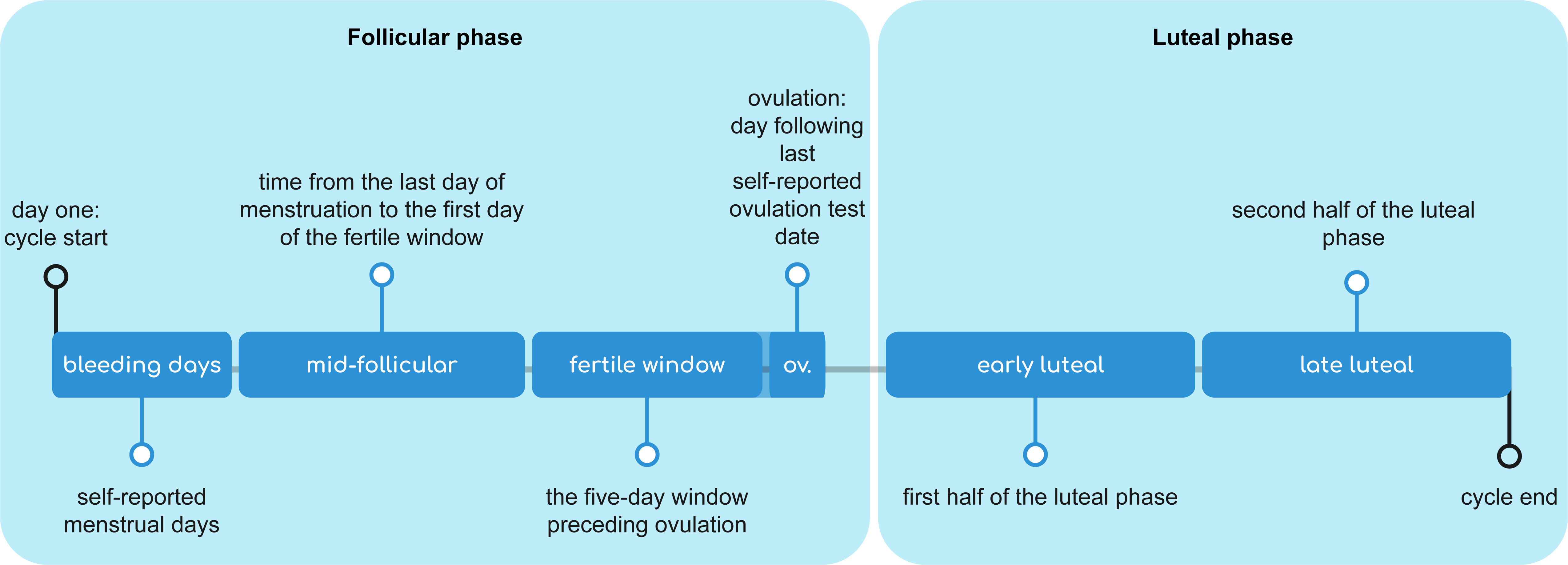

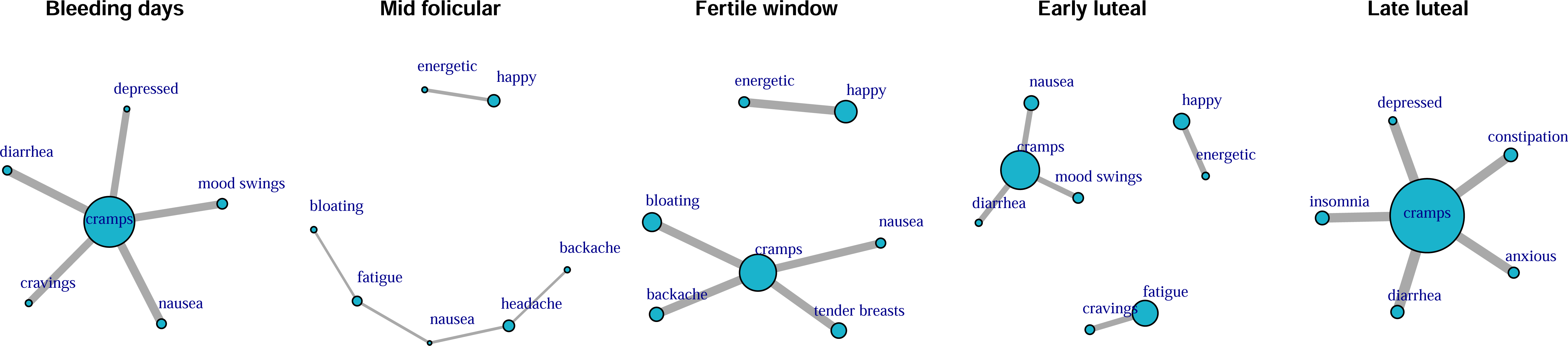

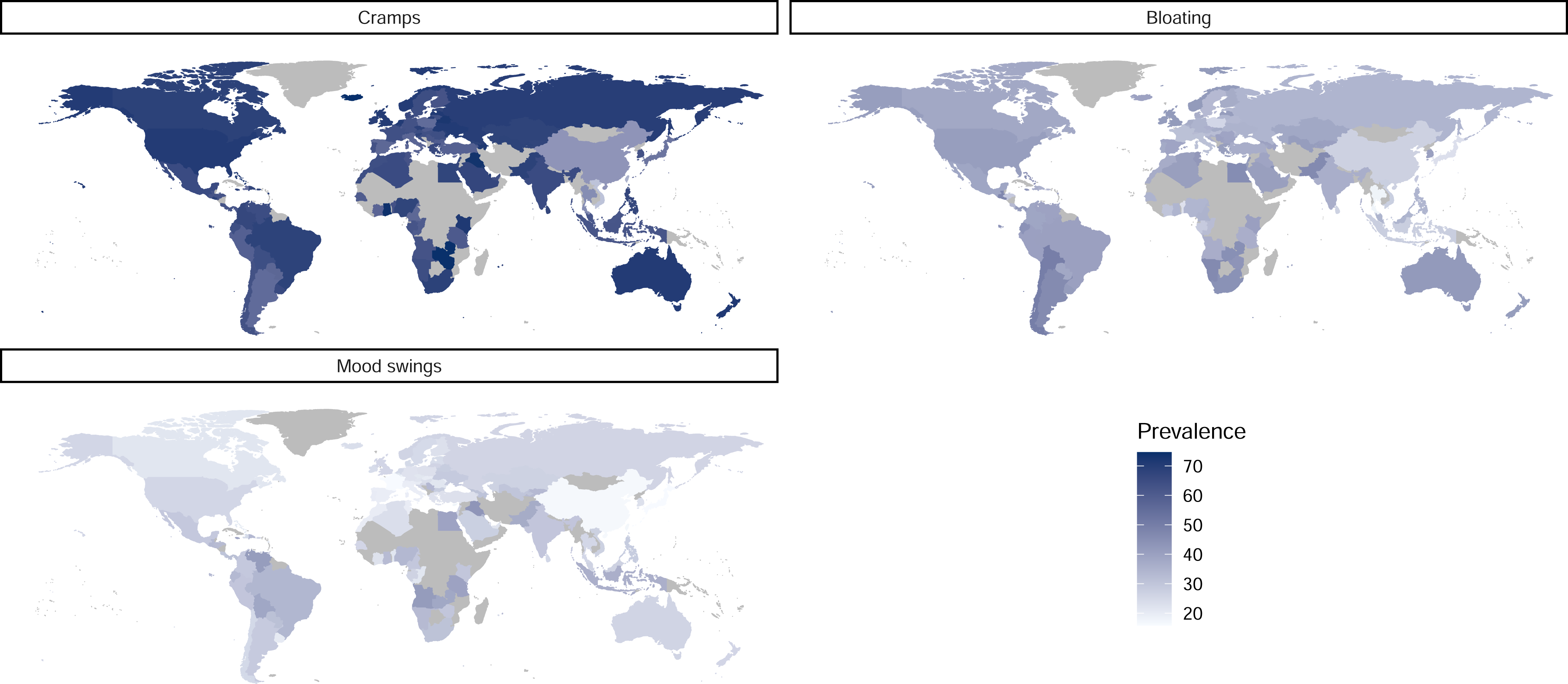

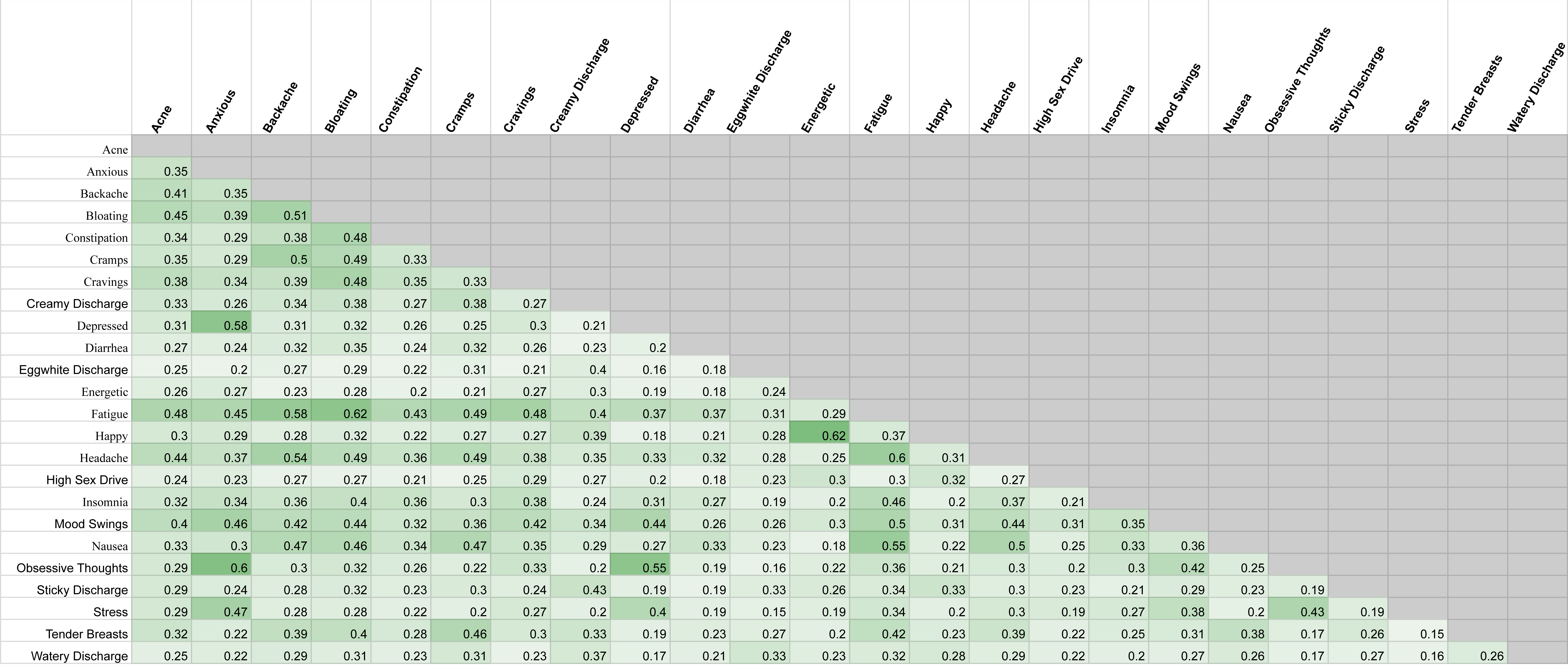

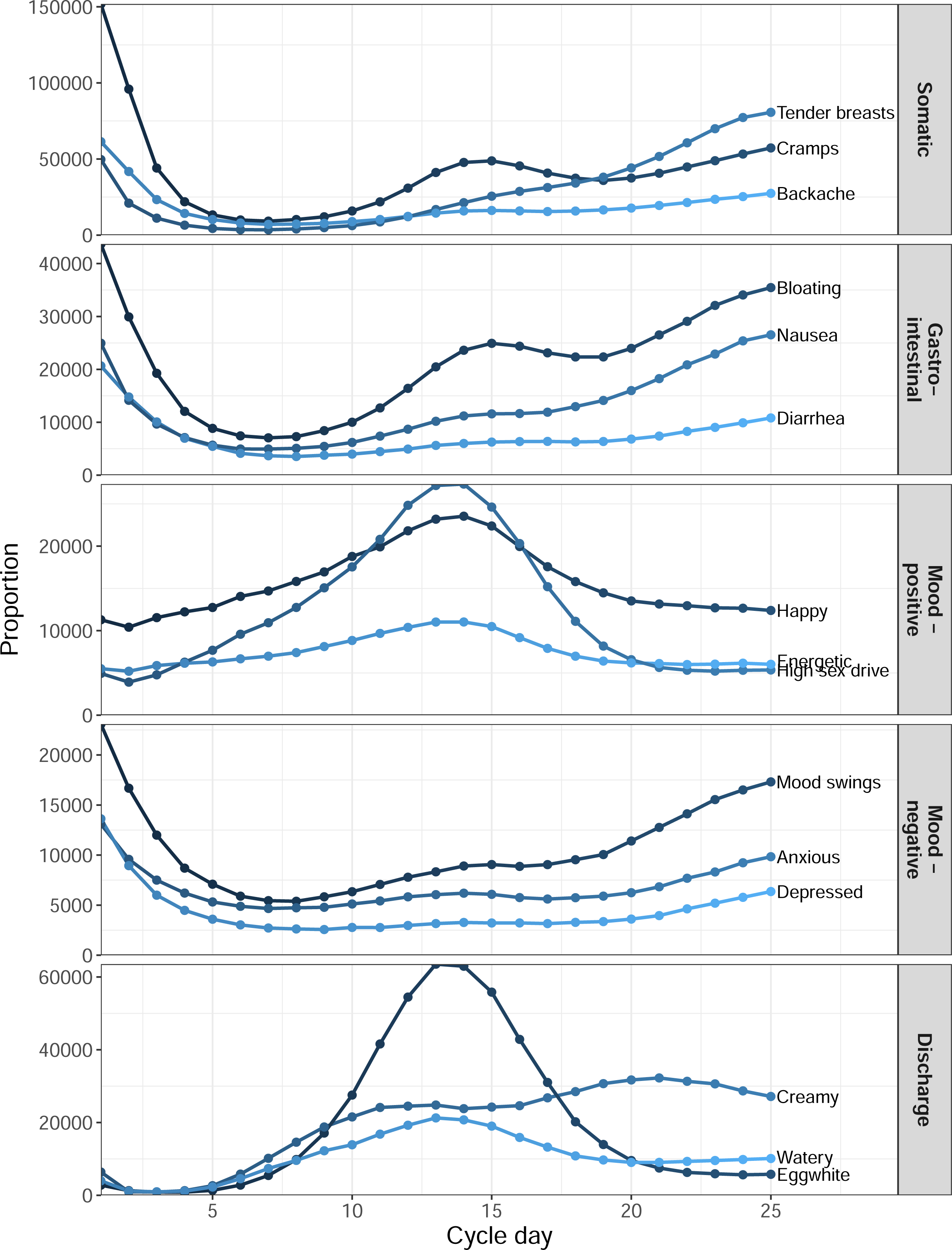

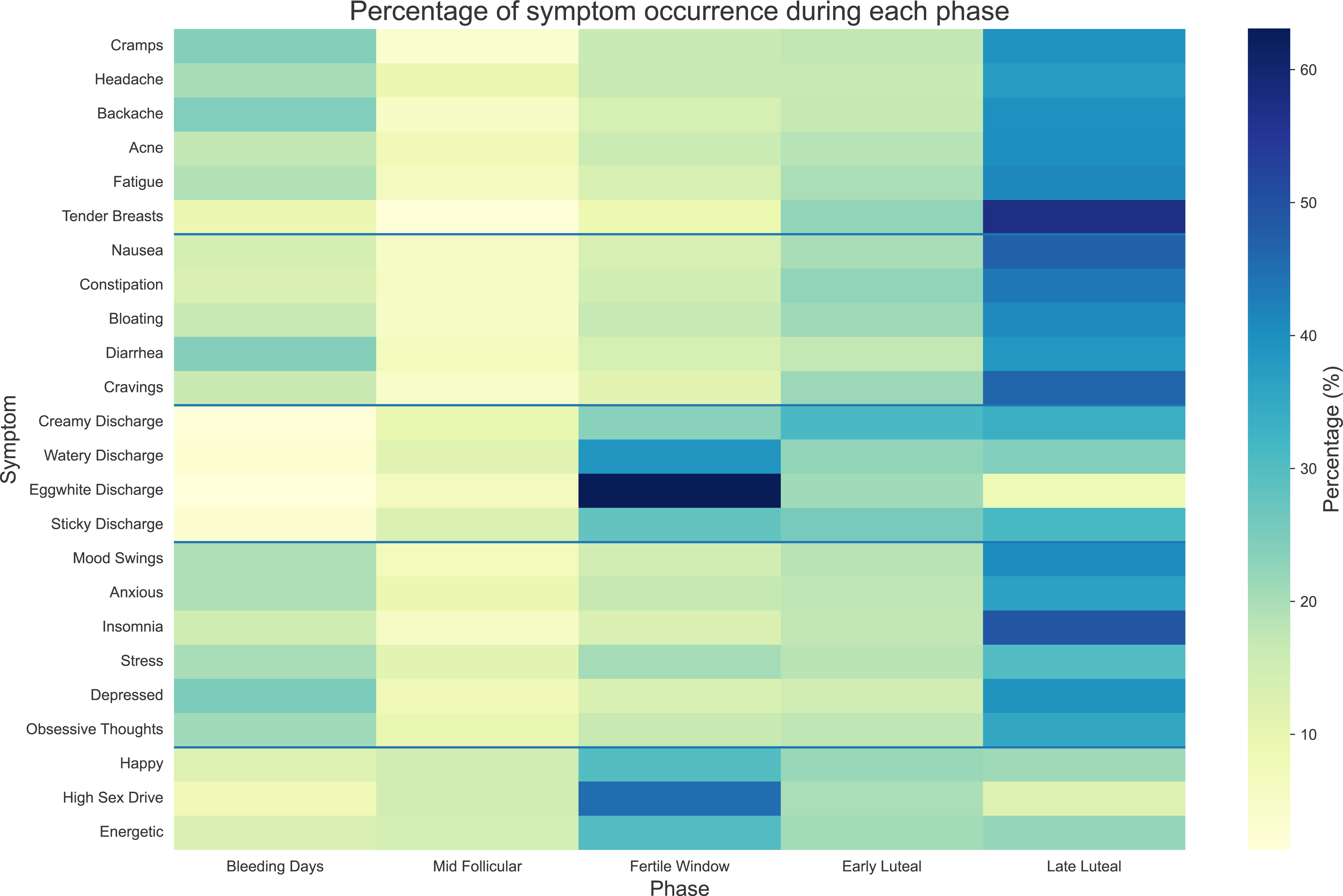

